# Predictions for COVID-19 Outbreak in India using epidemiological models

**DOI:** 10.1101/2020.04.02.20051466

**Authors:** Rajesh Ranjan

**Affiliations:** Department of Mechanical & Aerospace Engineering, The Ohio State University, Columbus, OH 43210

**Keywords:** COVID-19, Coronavirus, India, SARS-CoV-2, Epidemics, Social distancing

## Abstract

COVID-19 data from India is compared against several countries as well as key states in the US with a major outbreak, and it is found that the basic reproduction number *R*_0_ for India is in the expected range of 1.4-3.9. Further, the rate of growth of infections in India is very close to that in Washington and California. Exponential and classic susceptible-infected-recovered (SIR) models based on available data are used to make short and long-term predictions on a daily basis. Based on the SIR model, it is estimated that India will enter equilibrium by the end of May 2020 with the final epidemic size of approximately 13,000. However, this estimation will be invalid if India enters the stage of community transmission. The impact of social distancing, again with the assumption of no community transmission, is also assessed by comparing data from different geographical locations.

## 1 Introduction

Coronavirus disease 2019 (COVID-19) has presented an unprecedented challenge before the world. As of March 30, 2020, there have been about 0.8 million confirmed cases of COVID-2019 and about 40,000 reported deaths globally. About one-third of the world population is currently under lockdown to arrest the spread of this highly infectious disease. COVID-19 is caused by the novel coronavirus SARS-CoV-2, for which there is no specific medication or vaccine approved by medical authorities yet. This disease is transmitted by inhalation or contact with infected droplets or fomites, and the incubation period may range from 2 to 14 days[1]. Though the overall fatality rate is estimated to range from 2 to 3%, the disease can be fatal to elderly people (about 27% for 60+ age groups) and those with an underlying medical history. Studies[2, 1] have presented characteristics of COVID-19 disease describing symptoms and the latest developments.

India first reported a COVID-19 case in a student who returned from Wuhan, China on January 30, 2020. For future references, 2020 is the default year for all the dates, unless mentioned otherwise. Since then, there has been a gradual rise in the number of infections with 1,251 cases on March 30, among which there are 1,117 active cases, 102 recovered cases and 32 deaths. In response, India has implemented international travel bans and a strict lockdown. However, countries like India are at a greater risk because of a very large population density, limited infrastructure and healthcare systems to cater to very large demands. On the other hand, factors like warmer climate as well as humidity[3, 4], a large proportion of the young population, and possible immunity due to BCG vaccinations[5], may favor India. Most of these studies are preliminary and correlation-based, and therefore more evidence is required for arriving at concrete conclusions[6].

We hereby present epidemiological models for the spread of COVID-19 in India. Most of pandemics follow an exponential curve during the initial spread and eventually flatten out[7]. The current models are thus based on an exponential fit and logistic regression for short term and long term predictions, respectively. Further, in the latter case, Susceptible-Infectious-Recovered (SIR) compartment model[8] is used to include considerations for susceptibles, infectious, and recovered or deceased individuals. These models have shown a significant predictive ability for the growth of COVID-19 in India on a day-to-day basis so far.

We have also considered the possible effects of social distancing on the growth of infections. India announced a countrywide lockdown on March 24 for 21 days although a study by Singh and Adhikari[9] have suggested that this period may be insufficient for controlling the COVID-19 pandemic. In the present study, we assessed the effects of social distancing measures from the time of spread, by comparing lockdown status of India with several other countries, and states in the US and China. It is shown that the early action of lockdown in India, compared to many other countries/states, can have a favourable effect in limiting the final epidemic size. However, social and economical issues in a country of 1.34 billion population with high density pose significant challenges in enforcing strict social distancing.

As the current model is fully dependent on data, it is imperative to comment on the nature of this data. Different countries have different strategies for conducting COVID-19 diagnostic tests. In India, testing has largely been limited to individuals travelling from high-risk countries and their immediate contacts, as well as selected Pneumonia patients and symptomatic healthcare workers. As of March 30, India has tested 42,788 samples. Indian medical authorities have justified this strategy by testing randomly collected samples. Sahasranaman and Kumar[10] have compared basic reproduction number *R*_0_ from India and the world to analyze this strategy. *R*_0_ is the transmission rate given that the population has no immunity from past exposures or vaccination, nor any deliberate intervention in disease transmission. The number of infections grow and spread in the population if *R*_0_ *>* 1. They have found that *R*_0_ from India (*R*_0_ ≈ 0.43) is much smaller than the rest of the world (1.5 *< R*_0_ *<* 2.5), and the numbers reported in India may not be reflective of the actual number of cases. However, by considering a longer date range from March 10 to March 30, we show that the value of *R*_0_ from India is comparable to infection rates reported elsewhere. Further, we find that the growth of infections in India is comparable to that in California and Washington. In any case, current models do not depend on the testing strategy provided the same protocol is used throughout the time-period. In other words, the ratio of actual to reported number of cases will remain the same at any given time. On the other hand, the uncertainty due to exclusion of asymptomatic cases can be a major limitation in predictions with these models.

The current model does not account for factors such as the weather or humidity changes. Several studies[11] have reported that the efficacy of coronavirus may change when the weather becomes warmer. Other factors, such as differential immunity of Indians due to BCG vaccine[5] are already implicitly assumed in the data in the form of basic reproduction number.

Current models predict the transmissions due to stage-1 (individuals with a travel history to high-risk countries) and stage-2 (person-to-person contact). It should be noted that if the reported number of cases begins to exceed the predicted end-state systematically, then the pandemic will enter a new stage, and none of the models described above will be applicable. Although as of March 30, there is no strong evidence for community transmission (which can be achieved only by extensive testing and enlarging testing criteria). However, high population density as well as social and demographical issues puts India on a high risk for community transmission (stage-3). Although, the social distancing and rigorous contact tracing measures taken by India may help in containing these transmissions to small clusters, migration of labourers and workers could worsen the situation. Therefore, these factors should be considered while making informed decisions based on the present study.

The missing data of imported infections[12] may also affect the predictions but this may be a moot point as discussed below. India implemented a travel ban on March 12. The total number of air travellers who entered India in 2 months before this ban was around 1.5 million, and a gap between those being monitored for COVID-19 and total arrivals during this period has been reported[12]. Although if a significant number of infections were missed, a spike would have already appeared in the curve by March 30. It takes 2-14 days for the COVID-19 symptoms to appear, but no unusual events are noticed 18 days after the travel ban.

## 2 Epidemiological Models

### 2.1 Exponential Model

It is known that during early stages of a pandemic, the growth in the number of diagnosed infections with time is exponential. Thus, if the number of diagnosed infections *I*(*t*) over time *t* is known, one can find the growth rate *r*,

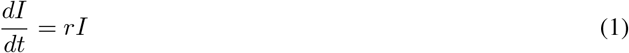

Integrating, we get-

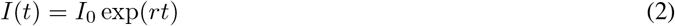

Where *I*_0_ is constant that can be obtained by fitting the curve with the available data. In this model, *I* is taken as the cumulative number of diagnoses (including individuals who have recovered or deceased).

### 2.2 Logistic Model

While the exponential model predicts the initial growth of a pandemic, it does not account for eventual decay and flattening out of curve. Logistic model on the other hand predicts the eventual decay but may fail in initial stages. In Logistic model, the growth is given by[13]:

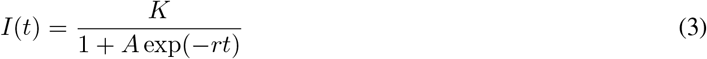

where *A* = (*K/I*_0_) − 1, assuming *K* ≫ *I*_0_ and hence *A* ≫ 1. For small time *t*, this approximates the exponential model.

### 2.3 SIR Model

Susceptible-Infectious-Recovered (SIR) model is a compartmental model that accounts for number of susceptibles *S*, number of infectious *I*, and the number of recovered or deceased (or immune) individuals *R*. Their distributions can be given as follows[8]:

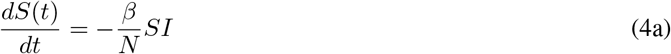

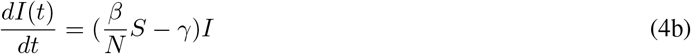

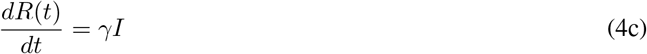

where *β* is the transmission rate, and *γ* is the average recovery rate. Note that here constant *N* is not the population of the country but population composed of susceptibles (*S*), infected (*I*) and recovered (*R*). In the present model, at any time the total population *N* = *S* + *I* + *R* remains constant as 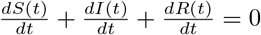 Initially, *N* is approximately equal to *S* as *I* is very small. In an outbreak, typically *N* will increase every day because more people may get affected due to local outbreaks. However, with quarantine and isolation, this number will slowly become constant. Hence, this SIR model, where *N* is assumed constant, is valid provided measures are taken to ensure *N* does not increase much with time. In the context of India, this model may be useful as rigorous social distancing measure has been introduced at an early stage and thus increase in *N* may not be significant. This model assumes equally likely recovery of everyone affected and hence does not consider factors like age etc.

The basic reproduction number in this model is given by-

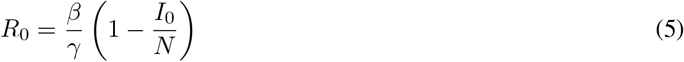

We assume a disease-free equilibrium (DFE) for a completely susceptible population i.e. the final number of infected people is zero. In the current approach, the initial guess of *γ* and *β* are obtained by setting *R*(0) = 0 and then the SIR equations are solved. Details of the implementation can be found in Batista[14]. For the COVID-19 predictions in China, Batista[14] has shown that both logistic and SIR give similar results. Hence, in this study only SIR results are included. For this purpose, the *fitViruscv19v3* code developed by McGee[15] is used.

## 3 Results

The data for the models have been taken from ‘Johns Hopkins University Coronavirus Data Stream’ that combines World Health Organization (WHO) and Center for Disease Control and Prevention (CDC) case data. For the exponential model, the data between March 11 and March 23 were used, when the number of reported infections were 62 and 499 respectively. For the SIR model, a longer range is required to obtain a reasonable estimate, and hence data for 21 days were considered starting from March 10, which is designated as the seed value. On this date, there were 56 individuals who had contracted the virus, out of which 39 were travel-related (stage-1) and 17 were person-to-person (stage-2) transmissions. There is no confirmed report of community transmission as of March 30. Hence, we consider that models with these seed values will give a good estimate for stage-1 and 2 transmissions. Further, like the study of Singh and Adhikari[9], we consider all cases to be symptomatic as it is not easy to estimate the number of asymptomatic cases. This could significantly underestimate the actual numbers of cases.

Before showing the models for India, we compare the growth of infections with several different countries and states in the US. There have been several reports (see for example, [10]) that have indicated that the initial slow growth of infections in India could be an artifact of its testing strategy, where testing is limited to specific individuals travelling from high-risk countries and their immediate contacts. Figure 1 shows the growth of infections in India from 1 to 1000, along with other countries and states with a reasonable number of daily international travellers. These places have used different testing strategies to report the number of infections. The growth rate in India is much smaller than places like New York and New Jersey, where the spread is very fast and it took only 16-17 days to reach 1,000 cases. Italy and France took about 29 and 43 days respectively to reach the same figure. On the other hand, places like California and Washington took a relatively long time (about 55-58 days), which is similar to India that took 59 days. Further, the growth curve of India is very close to that of Washington. As India is 9 days behind Washington in outbreak history, this information could be very useful as one may look at the Washington data to make predictions for India.

**Figure 1:**
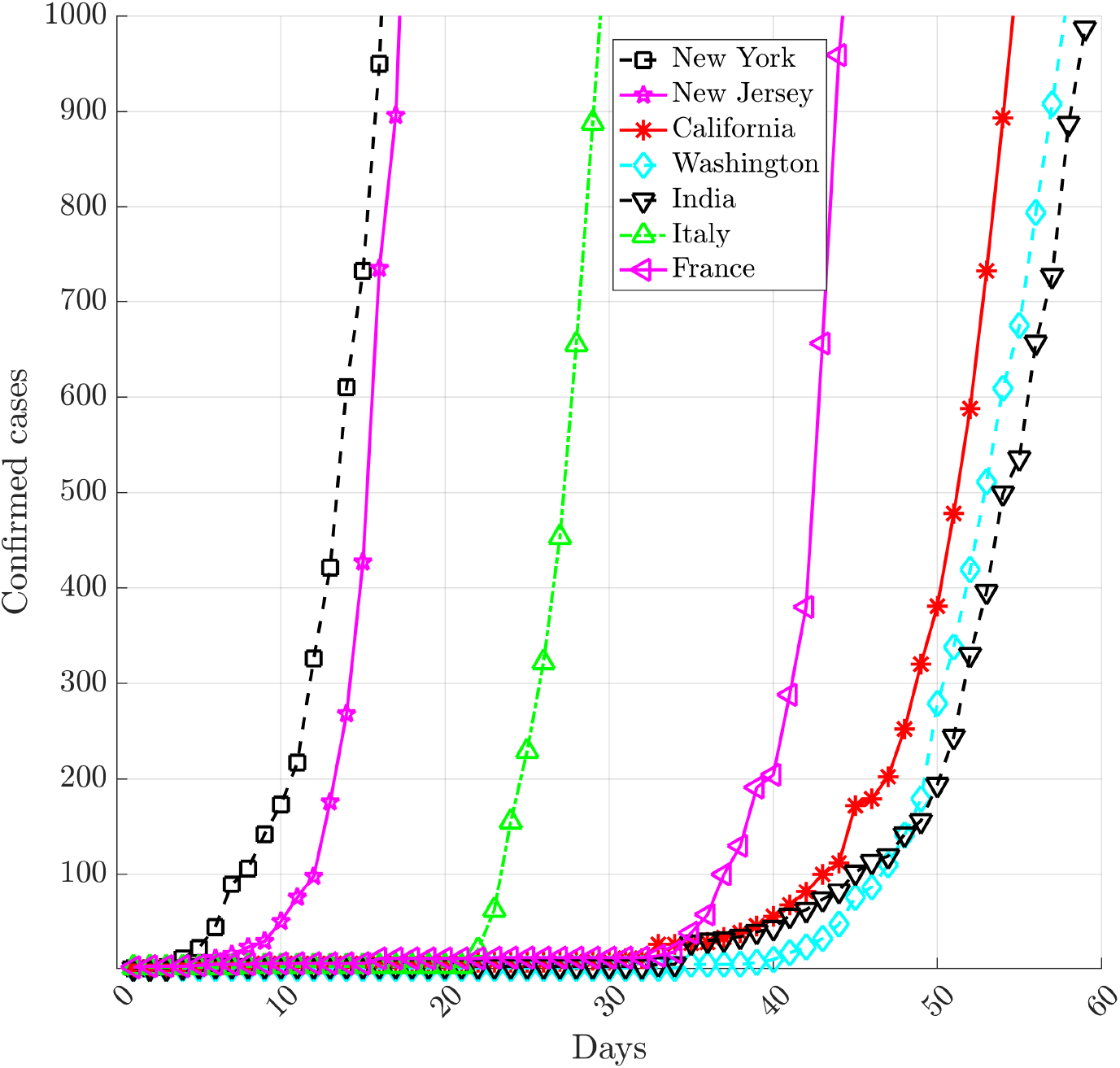
Days taken for infected cases to rise to 1000. The initial growth of epidemic in India is very close to that in Washington and California.

The results for the exponential and SIR mathematical models are discussed next. Figure 2 shows the exponential model for short and long term predictions. Table 1 shows the estimated coefficients for this model. The lower and upper bounds at 95% confidence level are given, along with other statistical parameters such as standard error, t-stat and p-value for each co-efficient. Statistical parameters of this regression model is shown in Table 2. The co-efficient of determination for this fit is *R*^2^ = 0.9768. Further, a very small p-value (∼ 0.000) indicate that all the regression parameters are statistically significant.

**Table 1:** Nonlinear regression model: *y* = *a*_1_ exp(*a*_2_*x*). Estimated Coefficients

|  | Estimate | 95% CI |  | SE | tStat | pValue |
| --- | --- | --- | --- | --- | --- | --- |
| $a_1$ | 39.64 | 29.91 | 49.37 | 4.3662 | 9.0788 | 3.8259e-06 |
| $a_2$ | 0.1887 | 0.1646 | 0.2127 | 0.010783 | 17.496 | 7.9017e-09 |

**Table 2:**
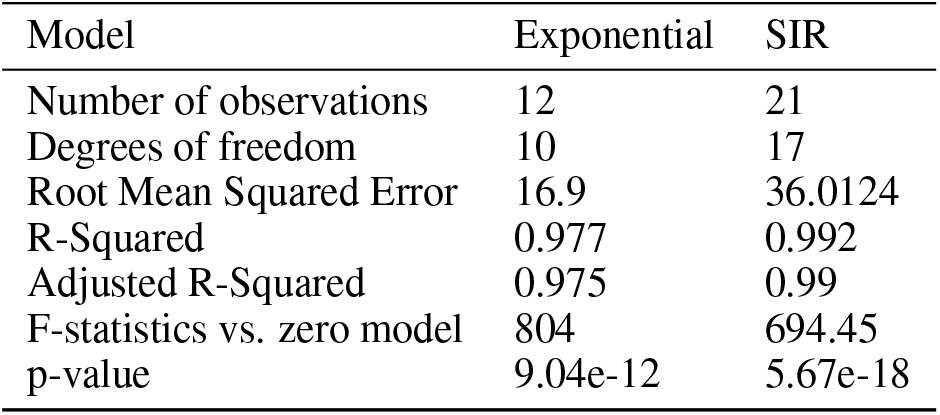
Statistics for Exponential and SIR model

| Model | Exponential | SIR |
| --- | --- | --- |
| Number of observations | 12 | 21 |
| Degrees of freedom | 10 | 17 |
| Root Mean Squared Error | 16.9 | 36.0124 |
| R-Squared | 0.977 | 0.992 |
| Adjusted R-Squared | 0.975 | 0.99 |
| F-statistics vs. zero model | 804 | 694.45 |
| p-value | 9.04e-12 | 5.67e-18 |

**Figure 2:**
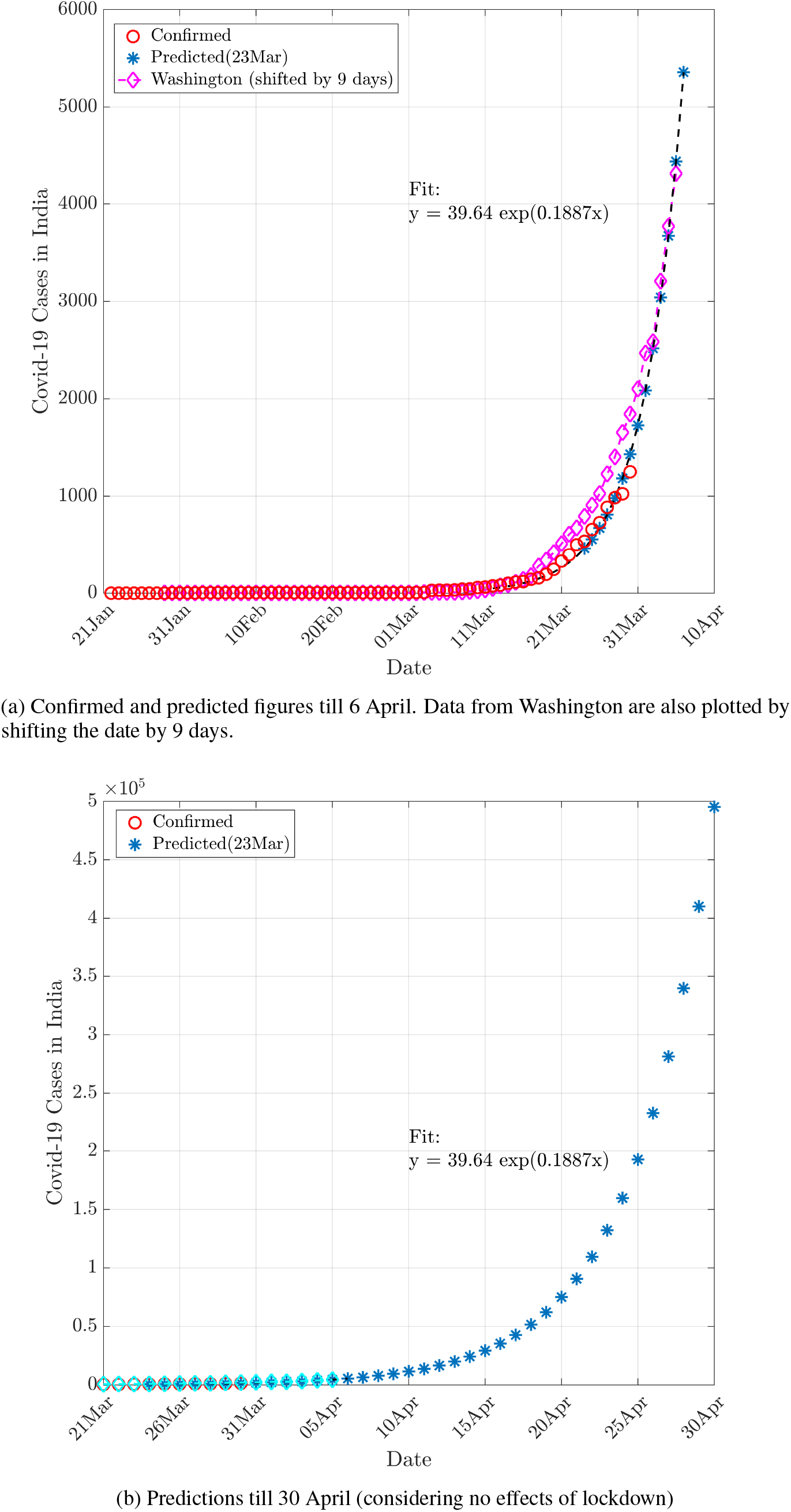
Predictions using the Exponential model. This model makes excellent short term predictions, and closely follow the Washington curve. The long-term predictions considering exponential growth present a worrying picture with about 0.5 million cases by end of April 2020.

Figure 2(a) shows confirmed as well as expected numbers of patients up to April 6. For comparison, the confirmed data from Washington are also shown. Though the model uses the data till March 23 for predictions, the number of reported cases are remarkably close to the number of predicted cases. According to this curve, India can have around 5,000 infected cases by April 6.

Though the exponential fit is not a good model for long term predictions, we present a pessimistic case by showing the results till April 30 using this model in Figure 2(b). It should be mentioned that these models do not consider any effect of social distancing due to lockdown that was enforced on March 24. Though, this presents a very frightening picture as the number of affected individuals may go up to 0.5 million by April 30, with a single-day increase of around 90,000 patients. However, this situation is going to be highly unlikely because of: (1) inability of this model to account for eventual decay in an epidemic, and (2) measures due to social distancing that result in flattening the curve.

Results for the SIR model, which has better abilities for long term forecasts, are shown in Figure 3. The statistical parameters for this regression model is also included in Table 2, with a coefficient of determination of *R*^2^ = 0.992, and p-value very close to zero indicating high statistical significance. As seen in the top panel of Figure 3, after the initial exponential phase till March 31, the acceleration phase starts and continues till April 13 when there is a peak of around 400 cases per day (bottom panel). The deceleration phase begins from this date and continues till April 30 after which asymptotic slow growth begins till flattening of the curve. The estimates from this model reaches a plateau at 12,416 cases, which is the final epidemic size. In the acceleration phase (April 1-April 13), the predictions from the SIR model differ from exponential model. For example, the exponential model suggests that the number of cases will cross 5,000 on April 6, but the prediction for the same using SIR model is on 12 April.

**Figure 3:**
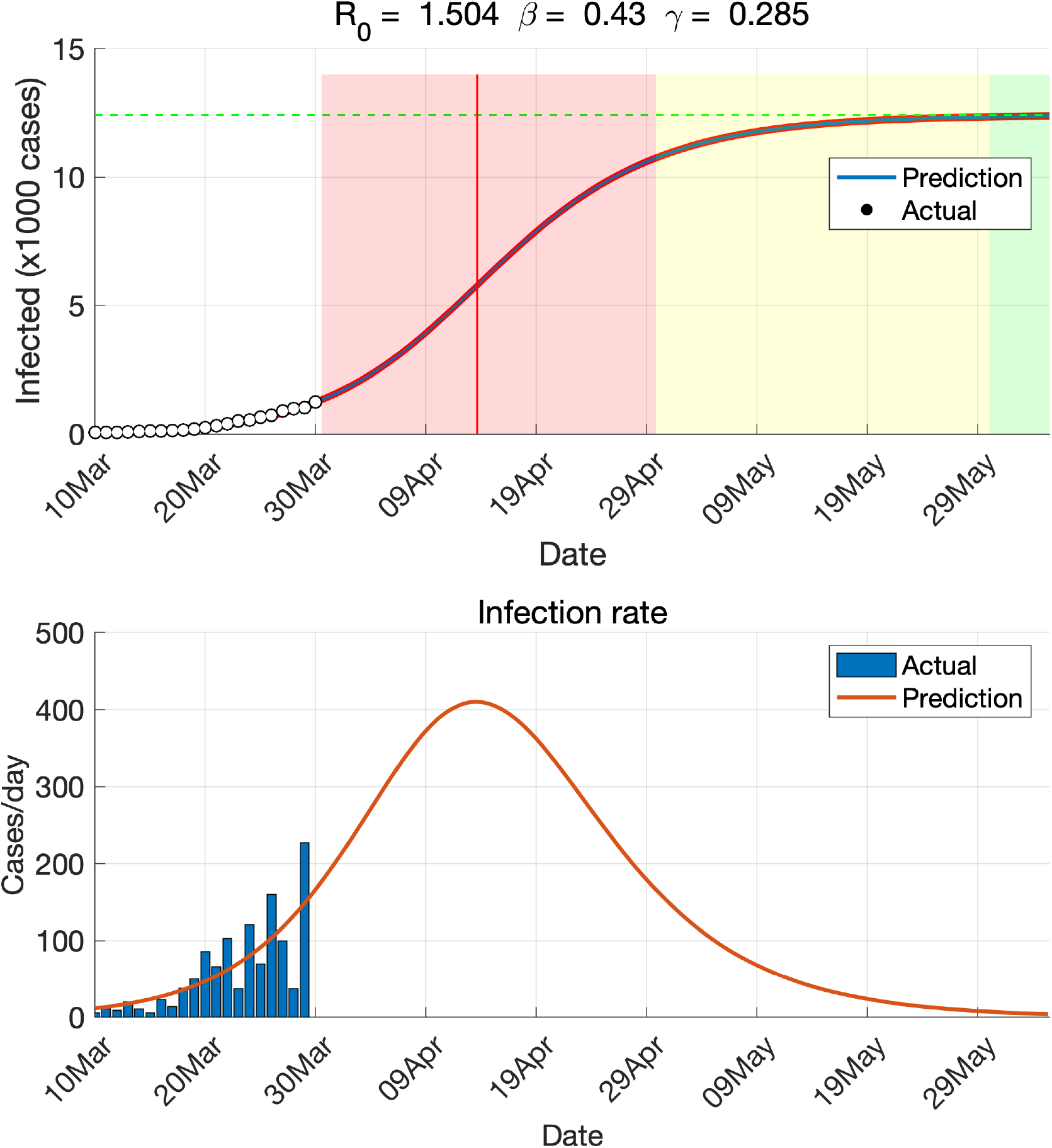
Predictions using the SIR model. This model gives final epidemic size around 13,000 that happens around the end of May 2020. Top panel shows different epidemic phases based on this model. White, red, yellow and green regions indicate initial exponential growth, fast growth (with positive and negative phase separated by red vertical line), asymptotic slow growth and curve flattening, respectively.

The basic reproduction number *R*_0_ using SIR model is 1.504, which is much higher than the initial values reported by Sahasranaman and Kumar[10] using the SIR model but it is close to the values reported by Deb and Majumdar[16] who used auto-regressive integrated moving average (ARIMA) model. The initial doubling time from the SIR model is 4.8 days. These values can be compared with estimated epidemiologic parameters from COVID-19 in Wuhan[17], where *R*_0_ and initial doubling time were about 1.9, and 5.4 days respectively. For US, particularly New York and New Jersey, the estimated *R*_0_ is very large[16]. However, as Ridenhour *et al*. [18] pointed that *R*_0_ “is a complicated property of an epidemic specific to the underlying model used to estimate it, the population being studied (in terms of contact patterns and demography), the host, the pathogen, and often the specific strain of the pathogen”. They suggested that this number may vary geographically due to changes in the environment, population structure, viral evolution, and immunity as well as healthcare and immigration policies.

Figure 3 suggests that the pandemic may reach equilibrium by end of May based on data till March 30. However, these estimations may change greatly if there is any jump or large departure in the trend. Further, as explained before, none of the models described above will be valid if India enters stage-3 (community transmission). On the other hand, the social distancing measures taken by India can help in arresting the growth much earlier. This aspect is analyzed in the next section.

### 4 Effects of Social Distancing

The effects of social distancing on COVID-19 outbreak have been studied by various researchers using different mathematical models [1, 19]. Here, instead of incorporating that effect into the mathematical model, we follow an empirical approach by learning from the data from different countries and states in the US.

It is known that the effects of social distancing become visible only after a few days from the lockdown. This is because the symptoms of the COVID-19 typically take some time to appear after getting infected from the Coronavirus SARS-CoV-2. The average incubation period is normally 5-6 days but it may take 2-14 days according to the CDC.

Table 3 shows the list of a few countries and cities with their lockdown status. Particularly, we focus on the lockdown in the Hubei province of China, where it was enforced on Feb 2. Figure 4 shows the number of confirmed cases as well as increase per day in Hubei. As shown in Figure 4(a), on the date of lockdown, Hubei had 11,177 cases (Lockdown in Wuhan, Capital city of Hubei was announced much earlier on January 23, when the country had just 830 cases).

**Table 3:** Lockdown status of different countries and states (On March 28, 2020)

| City/Country | Lockdown Date | Cases | Single day Peak<sup>2</sup> | Decline date | Remarks |
| --- | --- | --- | --- | --- | --- |
| India | 24-Mar | 536 | 160 |  | Police Enforced, Janta Curfew on 03/22 |
| Italy | 9-Mar | 9172 | 6557 |  | Police Enforced, Restricted domestic travel |
| Spain | 14-Mar | 6391 | 9630 |  | Police Enforced |
| France | 17-Mar | 7715 | 5141 |  | Strict nationwide lockdown, walk & travel restricted |
| Germany | 22-Mar | 24873 | 6933 |  | Strict social distancing measures |
| UK | 23-Mar | 43847 | 2769 |  | Police Enforced, domestic/local travel allowed |
| Hubei | 2-Feb | 11177 | 6200 | 14-Feb | Police enforced |
| New York | 22-Mar | 15168 | 7401 |  | New York State on PAUSE, domestic travel allowed |
| New Jersey | 21-Mar | 1327 | 2474 |  | Stay at Home, domestic travel allowed |
| Washington | 23-Mar | 2101 | 623 |  | Stay at Home, domestic travel allowed |
| California | 19-Mar | 952 | 877 |  | Stay at Home, domestic travel allowed |

**Figure 4:**
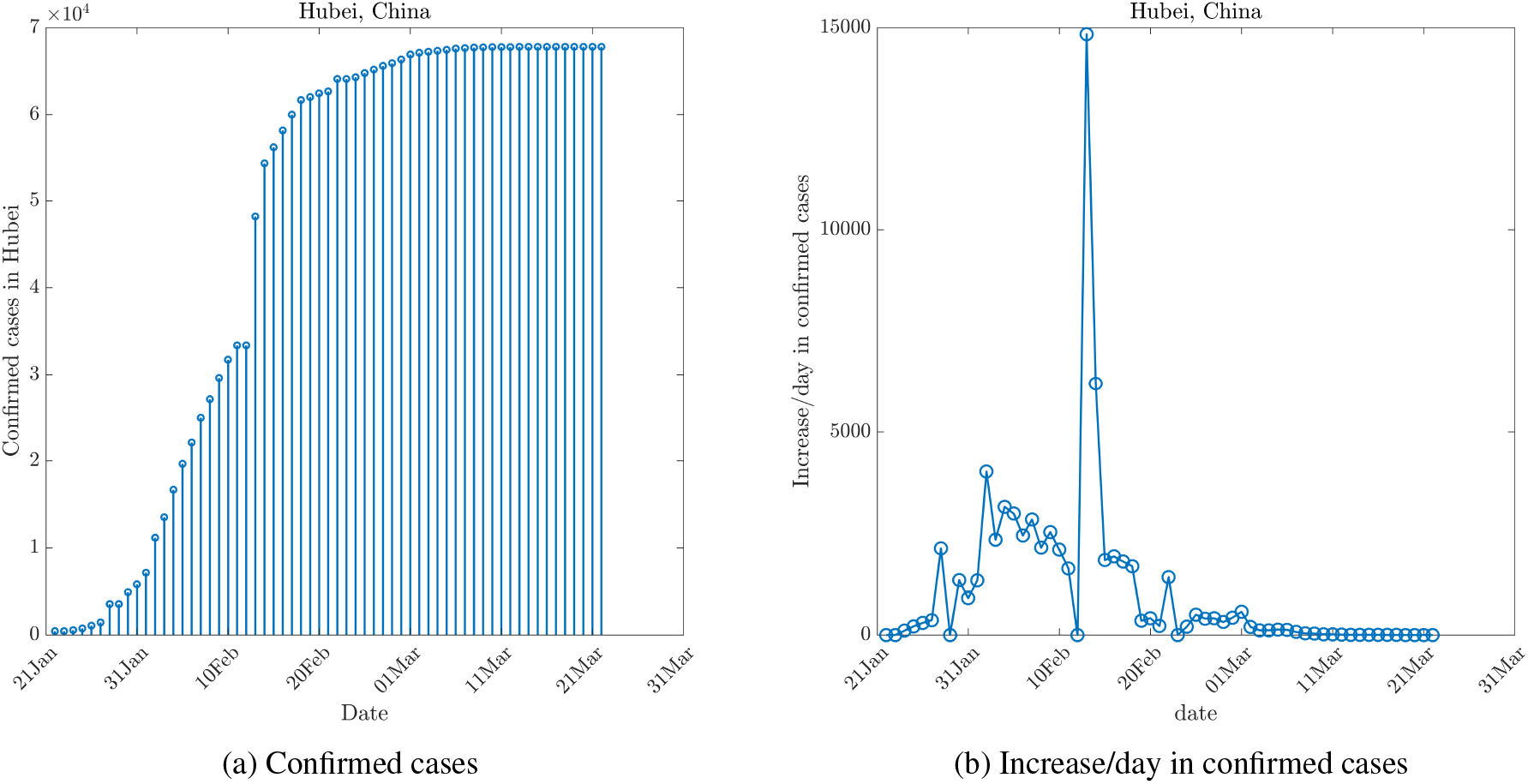
Effects of lockdown in Hubei, China. Lockdown in Hubei was enforced on Feb 2, 2020. However, the effects of lockdown in became visible only on Feb 14, 2020 as seen in (b), and the curve started flattening on Mar 4, 2020.

Figure 4(b) shows the increase in the number of cases per day. As seen, the cases in the Hubei province continued to rise till Feb 14 to reach a total of 54,406, with the total number of new cases reported on that day being about 6,200. It is to be noted that there is a slight anomaly in data with no new case being reported on Feb 14, whereas a sudden jump in the number of cases (∼ 15, 000) were reported on the following day. Since then, there was a gradual drop in reported numbers of cases, and on Feb 15 and Feb 21, the new cases reported were 1,843 and 220, respectively. From Feb 2 to Feb 15, it took around 14 days for the lockdown to show its effect. The flattening of the curve took further 15 days. i.e. from Feb 15 to March 2, as shown in Figure 4(a).

Another observation from Table 3 is that India was very early in enforcing lockdown when it had only 536 cases compared to many other countries/cities such as UK, Germany and New York where this strict measure was taken only after COVID-19 entered stage-3, and the spread became uncontrollable. This may become a key factor in controlling this pandemic in India.

Further, different countries have different norms as well as compliance levels due to practical considerations in enforcing the lockdown as shown in Remarks in Table 3. This may affect the final outcome. For example, the growth in Italy is still not stabilized after 19 days of lockdown. They have also seen the highest percentage of death as well. A large population over 60 years of age may be a primary factor. On the other hand, Spain has shown signs of decline from March 25 but it may be too early to confirm that. According to an estimate by Chang *et al*.[19] for COVID-19 outbreak in Australia, a reduction in incidence and prevalence can be visible only if the social distancing compliance levels exceed 80%.

Now, we focus on the impact of social distancing in India. India announced a strict lockdown on 24 March, when the number of reported cases was 536. Though on 22 March, India observed *Janta Curfew* (voluntary lockdown) and the public activity has been limited since that date.

Assuming the same pattern as Hubei in India as well (although the lockdown in Hubei was more stringent with strict police control over individual movements), we can assume that till April 8 (14 days from lockdown) very little effects of social distancing will be seen. By this date, India may have reported patients as many as 8000 (according to figure 2(b). This number may increase greatly if community transmission happens and spreads due to the movement of migrant workers and labourers.

However, after April 8, India should start seeing the effects of social distancing (provided it is enforced properly) and the curve should start flattening out. At its peak, India should expect around 1,500 patients on a single day (April 8) considering an exponential growth.

A recent study by Mandal *et al*.[20] has shown that social distancing can reduce cases by up to 62%. Assuming the uncertainty about the compliance in the enforcement of lockdown, we predict the social distancing effects with reductions of 30%, 50% and 70%. Figure 5 shows the impact of social distancing in these scenarios. Exponential growth is assumed throughout to account for the worst-case scenario. A reduction of 70% can bring the cases to a more controllable number i.e. a total of around 28,000 patients on April 20. Further, If India follows the case isolation strategy strictly, it is expected that this curve will start flattening out after that date (estimated based on the date from Hubei in Figure 4(a)).

**Figure 5:**
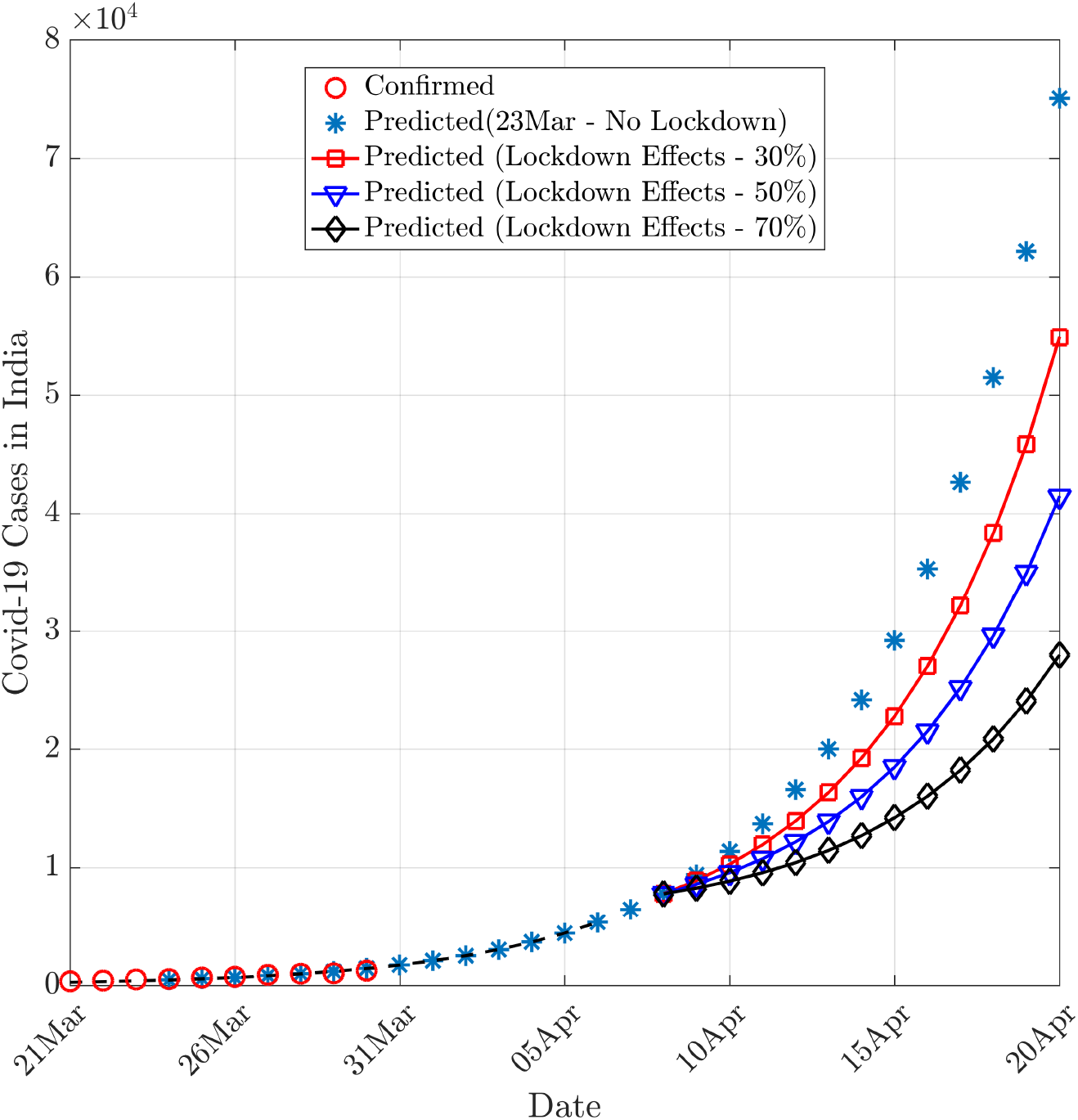
Effects of lockdown (assuming exponential growth throughout). Even with social distancing measures and 70% reduction in cases (most optimistic), there will be approximately 30,000 cases by April 20, 2020.

## 5 Conclusions

This exploratory evaluation shows that the transmission rate of COVID-19 in India is comparable to that in Washington state in the US. The curves describing the initial phase of the outbreak for both the locations are very close. As the beginning of the outbreak in Washington was about 9 days prior to that in India, the current data from the former can be used to make informative predictions for India. Along the similar argument, we show that the transmission rate and basic reproduction number *R*_0_ for India are in the expected range. This is contrary to several reports questioning the testing strategies adopted by India quoting a low transmission rate and hence the smaller basic reproduction number. Two epidemiological models - a simple exponential model and an SIR model, are used respectively to forecast short and long term outcomes. These models assume all the seed cases to be symptomatic, which may underestimate the actual numbers due to an uncertain number of asymptomatic individuals. With this limitation, the exponential model based on data till March 23 nicely predicts the values till today (March 30). The SIR model based on data till March 30 indicates that India will enter equilibrium by the end of May with an estimated total number of infected cases to be approximately 13,000. It is estimated that the impact of social distancing will be visible after April 8 following which we may see a significant reduction in the reported infections. However, India is on high risk to enter into community transmission due to reported violation of quarantine norms by individuals as well as other social and demographic issues. The predictions made using the current epidemiological models in the current work will be invalid if such an event occurs. Finally, the model is as good as the underlying data. Because of real time change in data daily, the predictions will accordingly change. Hence, the results from this paper should be used only for qualitative understanding and reasonable estimate of the nature of outbreak, but are not advisable for any decision making or policy change.

## Data Availability

The data that support the findings of this study are publicly available.

https://github.com/CSSEGISandData/COVID-19

## 6 Acknowledgment

The author would like to acknowledge Dr. Deepti Chugh (The Ohio State University) and Dr. Sudheendra N R Rao (Scientific Advisor, Organization for Rare Diseases India) for their useful inputs in this work.

## Footnotes

2 Data till March 28, 2020. Hubei has an anomaly in data as no new case was reported on Feb 12 and abruptly high number of cases (*∼* 15000) were reported on the following day.

